# Disentangling the drivers of heterogeneity in SARS-CoV-2 transmission from data on viral load and daily contact rates

**DOI:** 10.1101/2024.08.15.24311977

**Authors:** Billy J Quilty, Lloyd AC Chapman, James D Munday, Kerry LM Wong, Amy Gimma, Suzanne Pickering, Stuart JD Neil, Rui Pedro Galao, W John Edmunds, Christopher I Jarvis, Adam J Kucharski

## Abstract

SARS-CoV-2 transmission is highly overdispersed, with a minority of individuals responsible for the majority of transmission, though the drivers of this heterogeneity are unclear. Here, we assess the contribution of variation in viral load and daily contact rates to this heterogeneity by combining published viral load estimates and contact survey data in a mathematical model to estimate the secondary infection distribution. Using data from the BBC Pandemic and CoMix contact surveys, we estimate the secondary infection distribution throughout the pandemic in the UK in 2020, and the effectiveness of frequent and pre-event rapid testing for reducing superspreading events. We find that individual heterogeneity in contacts rather than individual heterogeneity in shedding is the main driver of observed heterogeneity in the secondary infection distribution. Our results suggest that everyone testing every 3 days would reduce the reproduction number below 1 and be equivalent in terms of impact on secondary infections to everyone testing only before events with a minimum event size of 10 for pre-pandemic contact levels. This work demonstrates the potential for using viral load and contact data to estimate heterogeneity in transmission and the effectiveness of rapid testing strategies for curbing transmission in future pandemics.

**Author Summary:** SARS-CoV-2 spreads mainly through superspreading, with around 20% of infected individuals responsible for around 80% of secondary infections. Previous studies have inferred this using plausible assumptions about contact rates and viral load dynamics. Here, we instead integrate data from real contact surveys conducted in the UK before and during the COVID-19 pandemic with published estimates of viral load trajectories and infectiousness to estimate the average and variation in numbers of secondary infections per case over the first year of the pandemic. Our results are consistent with observed reductions in secondary infections during periods of contact restrictions in the UK. We show that variation in numbers of daily contacts can be used to monitor superspreading risk in real-time during an epidemic. When combined with regular rapid testing, which identifies individuals with high viral loads when they are most infectious, this offers a means of effectively reducing transmission and avoiding costly blanket interventions by targeting restrictions to when and where they are most needed.

## Introduction

Transmission of SARS-CoV-2 occurs primarily through superspreading, with 20% of infections generating around 80% of secondary infections [1]. A review and meta-regression by Chen et al. [2] indicates that substantial variation in the respiratory viral load of individuals infected with SARS-CoV-2 is a driver of overdispersion in secondary infection generation. However, as most studies cited measured viral load at one point over the course of infection, this review could not answer whether this is due to some individuals being more infectious than others generally (“between-person infectiousness variability” hypothesis) or whether most individuals pass through a highly infectious period which happens to coincide with a period of high contact (“between-person contact variability and within-person infectiousness variability” hypothesis). High contact rates are a prerequisite for infecting a large number of people, and hence the potential for superspreading should have varied over the course of the COVID-19 pandemic as contact distributions changed with the enactment and relaxation of restrictions.

In this paper, we reconstruct the secondary infection distribution of SARS-CoV-2 using a model of intra- and inter-host heterogeneity in infectiousness derived from published data on viral load trajectories and infectivity combined with data on reported numbers of daily contacts from two social contact surveys in the UK. As overdispersion in the social contact data has not previously been analysed or reported, we first perform a descriptive analysis of the social contact data. While previous models of superspreading of SARS-CoV-2 have either fitted to summary data on key epidemiological metrics, such as the mean reproduction number and distributions of individual-level numbers of secondary cases from contact tracing studies [3], to estimate social contact rates, or considered a wide range of plausible contact rates [4], here we estimate the secondary infection distribution from first principles using data on social contacts gathered prior to and during the pandemic in the UK. This allows us to characterise variation in the secondary infection distribution over time under different levels of restrictions on contacts. We also consider the impact of rapid lateral flow antigen tests (LFTs), which are able to detect individuals with high viral loads when they are most likely to be infectious [5, 6], taken regularly or before events on the mean reproduction number and the potential for superspreading with differing levels of adherence and background contact rates.

The distribution of the number of secondary infections generated by each infectious individual *i*, *R_i_*, can be assumed to be negative binomial with mean equal to the mean number of secondary infections *R* and overdispersion parameter *k* representing the variation in the number of secondary infections, with smaller values of *k* representing greater variation,

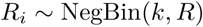

such that the variance in *R_i_* is

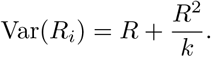

Even if the mean number of secondary infections *R* is below 1, there may still be a considerable probability of one or more secondary infections if *k* is small. We estimate the utility of regular rapid LFTs on reducing *R* and the potential for superspreading events by decreasing variation in numbers of secondary infections, that is, increasing *k*.

## Methods

### Ethics

This study only involved secondary analysis of publicly available contact survey data and SARS-CoV-2 viral load data. Secondary analysis of these data was approved by the London School of Hygiene and Tropical Medicine Observational Research Ethics Committee (ref 14400 and ref 21795).

### Contact data

#### BBC Pandemic survey

The BBC Pandemic contact survey was conducted between September 2017 and December 2018 as part of a BBC Four documentary and involved over 40,000 participants (full details are published elsewhere [7, 8, 9]). Participants used an app to record their personal basic demographic information and the number of social contacts they made during the previous 24-hour period, as well as information such as the contact’s age, type of interaction, and setting (home, work, school, other). Children under 13 years of age were excluded from participating for ethical reasons.

#### CoMix survey

The CoMix survey was a behavioural survey launched on 24th of March 2020 to gather social and behavioural data to aid the response to the COVID-19 pandemic. The contact survey was based on the POLYMOD contact survey [10]. The sample was broadly representative of the UK adult population. Participants were invited to respond to the survey once every two weeks. Weekly data was collected by running two alternating panels. Parents completed the survey on behalf of children (17 years old or younger). Participants recorded direct, face-to-face contacts made on the previous day, specifying certain characteristics for each contact including the age and sex of the contact, whether contact was physical (skin-to-skin contact), and where contact occurred (e.g. at home, work, while undertaking leisure activities, etc.). Full details have been published elsewhere [11].

From the 24th May 2020 onwards, the CoMix survey included the ability to record an estimated mass contacts count in situations where it would be infeasible to record the detailed information of all contacts made, such as large gatherings. Since the BBC Pandemic survey did not include this option, and this potentially truncated the true contact distribution, we conducted a sensitivity analysis of the effect on *R* and *k* of imputing a heavier tail for the BBC Pandemic contact distribution. We did this by fitting a generalised Pareto distribution to the number of individuals reporting over 250 contacts in relaxed restrictions time periods (“Relaxed restrictions” and “School reopening” in Table 1) from the CoMix data and sampling proportionally from this distribution for the BBC Pandemic contacts.

**Table 1.**
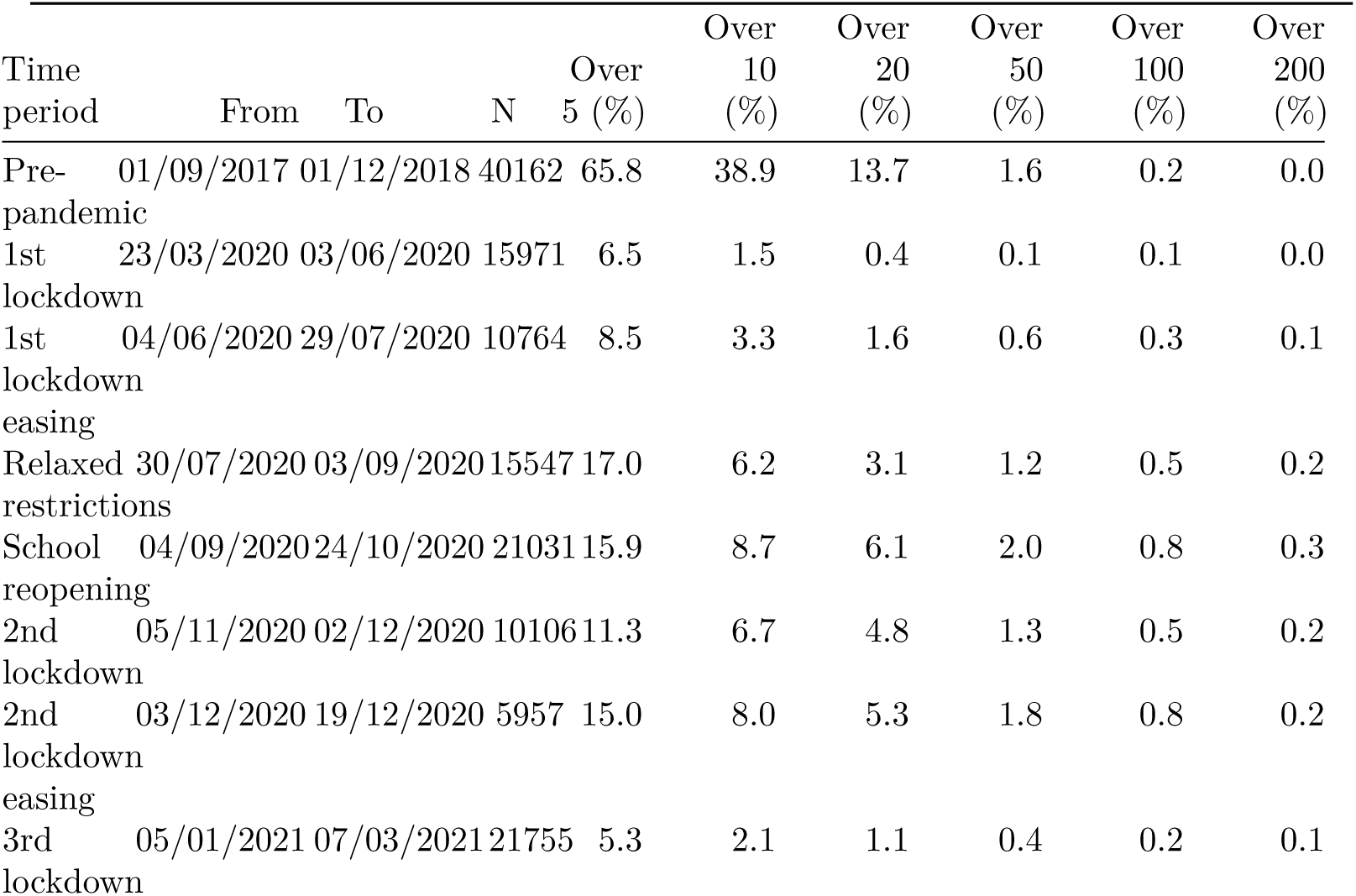
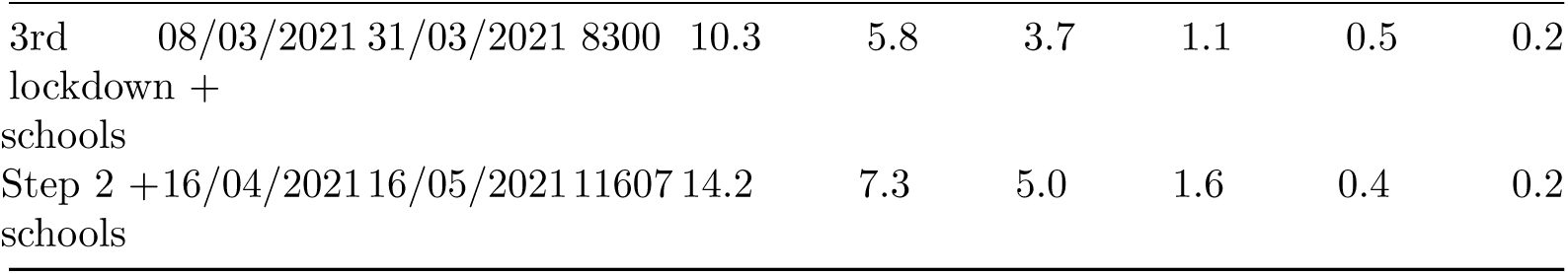
Percentage of participants in the CoMix and BBC Pandemic surveys with more than a certain number of contacts in different time periods in the UK from March 2020 to May 2021, and pre-pandemic in 2017-2018.

### Analysis of social contact data

We compared contact distributions (overall and stratified by household and out-of-household) for the BBC Pandemic survey [7, 8, 9] and CoMix survey [12, 11]. To account for differences in numbers of participants per time period, we calculated and plotted the percentage of participants reporting more than 5, 10, 20, 50, 100, and 200 contacts in the 24-hour period prior to filling in the survey for the BBC Pandemic survey (here referred to as Pre-pandemic) and nine indicative time periods between March 2020 and May 2021 from the CoMix survey representing different levels of restrictions, as defined in previous work [12, 11] (Table 1). We fitted negative binomial distributions to numbers of daily contacts in the different time periods to estimate the mean and dispersion of the contact distributions. To assess whether the negative binomial adequately captured the contact distribution, or whether an excess of zero-contact individuals required a zero-inflated extension, we additionally fitted a zero-inflated negative binomial (ZINB) distribution to the per-individual daily contact counts for three indicative time periods (Pre-pandemic, 1st lockdown, School reopening) and compared models using the Akaike information criterion (AIC) and a likelihood ratio test (LRT). The negative binomial was found to be sufficient in all periods (zero-inflation parameter *π <* 6 *×* 10*^−^*^8^, ΔAIC = +2.0, LRT *p >* 0.97; Table S2).

### Reconstruction of secondary infection distribution

We reconstructed the secondary infection distribution of SARS-CoV-2 in the UK over the course of 2020 as described below. We restricted our analysis to the first six time periods of the CoMix survey in 2020 (23rd March to 19th December) prior to the widespread emergence of more transmissible variants and mass vaccination.

#### Viral load trajectories

To predict the distribution of individual-level transmission at different points in time, we simulated 10,000 individual respiratory viral load trajectories of index cases over the course of infection. The viral load of each index case was constructed from a piecewise linear function of their Ct values defined by a proliferation phase (days from exposure to peak, *d_p_*), a clearance phase (days from peak to cessation, *d_r_*) and a peak Ct value, Ct*_p_*. The three parameters were drawn from distributions given in Kissler et al. [13] (Figure 1):

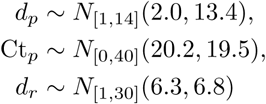

**Figure 1.**
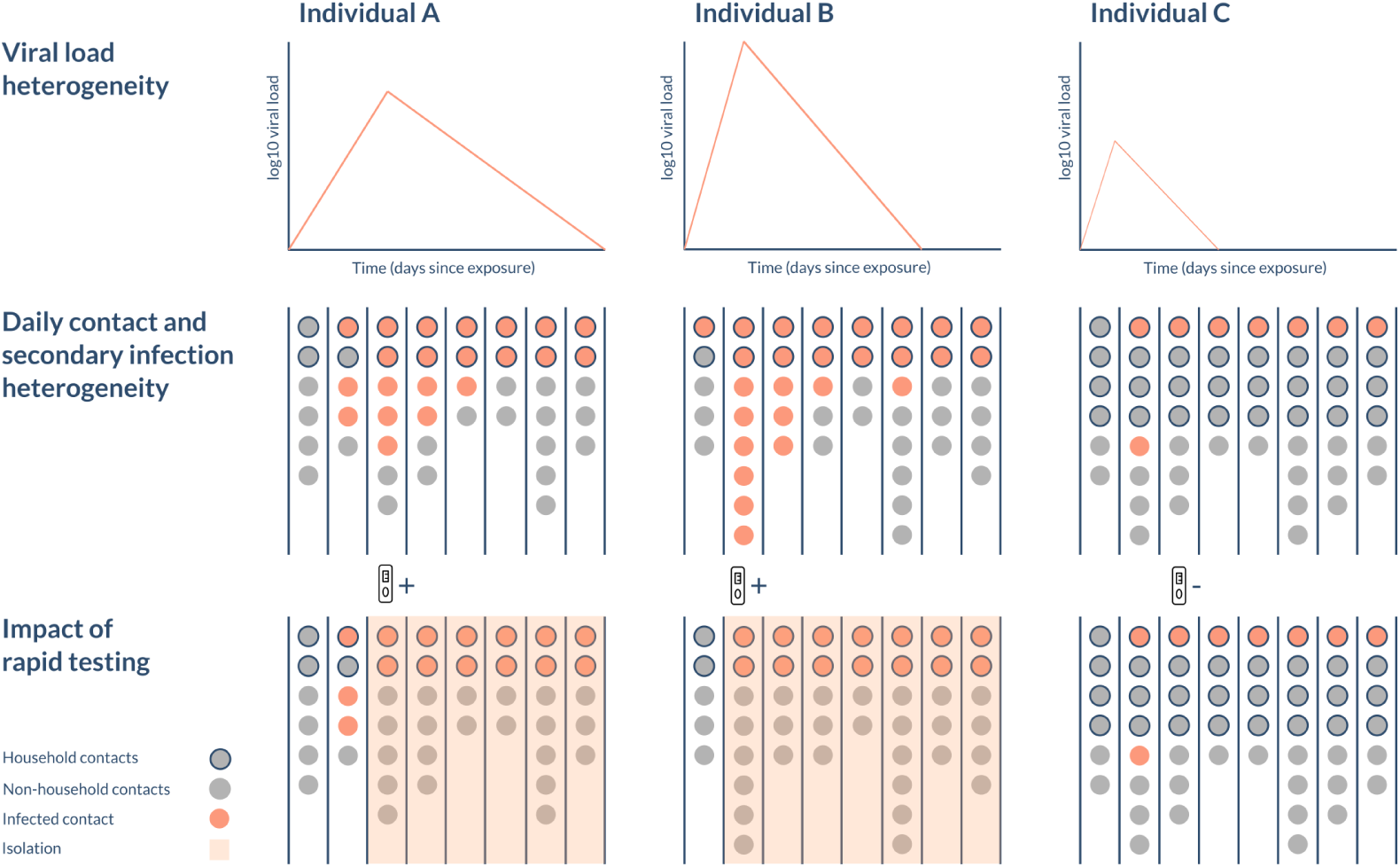
Schematic of the model. The viral load and number of daily contacts (circles) varies from person to person and over time, influencing the number of secondary infections (stratified by household (outlined) and non-household (no outline)) they generate. The viral load at time of testing also determines the likelihood they will test positive and subsequently isolate.

where *N*_[_*_a,b_*_]_(*µ, σ*^2^) denotes a truncated normal distribution with mean *µ* and variance *σ*^2^, and lower and upper limits *a* and *b*. The Ct value at exposure and cessation of shedding was assumed to be 40 (negative).

The Ct value was converted to viral load vl (in log10 RNA copies/ml) via the following formula [13]:

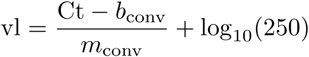

where *m*_conv_ = *−*3.60971 and *b*_conv_ = 40.93733 are the slope and intercept of a linear regression of Ct value on log10-transformed RNA concentration from a standard curve.

#### Infectiousness

The probability of infectiousness for a given viral load (in Ct) was estimated by fitting a logistic regression model to the probability of culturing virus at that viral load [14] (Figure 3B), *p*_culture_:

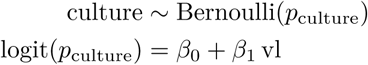

where culture is the binary outcome of whether or not virus was cultured. The probability of infectiousness over the course of each individual’s viral load trajectory was then calculated using the estimated regression coefficients *β*_0_ and *β*_1_ to produce infectiousness trajectories (Figure 3C). Uncertainty in *β*_0_ and *β*_1_ was accounted for by drawing values for each individual from a bivariate normal distribution centred at the maximum likelihood estimate *β*^ = (*β*^_0_*, β*^_1_) = (*−*14.8, 2.1):

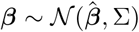

where Σ is the estimated covariance matrix from the logistic regression. Drawing values for the regression coefficients in this way may be taken to represent a degree of variation in infectiousness between individuals for the same viral load, as Ke et al. [6] concluded was necessary to explain observed heterogeneity in infectiousness. We use viral culture positivity as our proxy for infectiousness because it indicates the presence of replication-competent virus and is therefore believed to provide a better indicator of transmission potential than Ct values from PCR measurements [15]. It also allows us to make use of empirical relationships linking viral load, viral culture positivity, and test detection: (i) viral load determines the probability of culturing viable virus, with the logistic relationship fitted directly from data in Pickering et al. [14] (Figure 3B); and (ii) LFT positivity closely tracks viable virus in controlled human challenge studies [5].

#### Contacts and contact durations

For the pre-pandemic time period and each of the pandemic time periods, we randomly sampled the number of household contacts *N*_HH_*_,i_* and daily non-household contacts *N*_NHH_*_,i,t_* for index case *i*, where *t* is the time since exposure, from the empirical contact distribution for that time period from the BBC Pandemic contact survey [7, 8, 9] and CoMix contact survey [12, 11]. The number of household and non-household contacts were sampled jointly by sampling a survey participant at random, to preserve correlation in individual-level numbers of household and non-household contacts. The numbers of non-household contacts on different days were sampled from the repeat survey responses of the chosen participant for the given time period, to maintain correlation in individual-level numbers of non-household contacts over time. Each household and non-household contact had a duration, dur_HH_*_,i,j,t_* and dur_NHH_*_,i,j,t_*, defined as the proportion of a 24-hour period that the contact lasted (Figure S1). For the pandemic time periods the duration was sampled from the contact durations for the same survey participant as the contact numbers, where these were available. This ensured that correlation between number of contacts and contact duration was maintained (Figure S2). For the pre-pandemic time period, the duration was sampled from the contact durations in the POLYMOD survey, as contact duration data is not publicly available for the BBC Pandemic survey. Where the contact duration was missing, it was randomly sampled from the pre-pandemic or pandemic contact duration distribution according to whether it was a pre-pandemic or pandemic contact.

#### Infections

The infection process for contact between index case *i* and their contact *j* was modelled as Bernoulli for both household and non-household infections

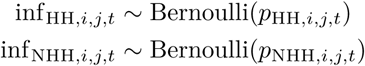

with the probability of *i* infecting *j* on day *t* in location X, *p*_X*,i,j,t*_, where X *∈ {*HH, NHH*}*, given by

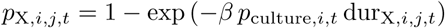

where *β* is a scaling parameter which converts the product of the relative infectiousness of *i* on the day of contact, *p*_culture_*_,i,t_*, and the duration of contact, dur_X_*_,i,j,t_*, into a transmission rate. This process was iterated over each day of the index case’s infection up to a maximum infection duration of 44 days in the baseline analysis. While we modelled infection events for household members on each day of the index case’s infection, we only counted the first infection that occurred for each household member. In other words, household members could only be infected once. We assumed uniform susceptibility of individuals in the model, which does not vary by, for example, age.

The number of secondary infections for index case *i* (i.e. their individual-level reproduction number), *R_i_*, was therefore given by:

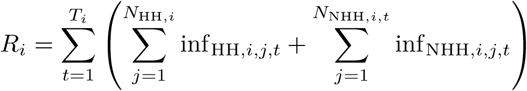

where *T_i_* is index case *i*’s duration of infection (defined as having infection detectable by PCR). We then estimated the mean, *R*, and overdispersion, *k*, of the number of secondary infections by fitting a negative binomial distribution to the *R_i_* values across all index cases. 95% percentile bootstrap confidence intervals (CIs) for the *R* and *k* estimates were calculated by bootstrapping the simulated numbers of secondary infections with 1000 bootstrap samples, calculating *R* and *k* for each bootstrap sample and then finding the 2.5 and 97.5 percentiles of the distributions of the bootstrap estimates. We calibrated *β* such that the basic reproduction number *R*_0_, the reproduction number for the pre-pandemic time period, was 2.5, based on estimates from the literature [12, 16, 17], for the baseline analysis and all sensitivity analyses varying heterogeneity in contact rates and viral load. We obtained values of *β* between 1.1 and 4.7.

### Contribution of contact and viral load heterogeneity to heterogeneity in secondary infections

To investigate the contribution of heterogeneity in contact rates and heterogeneity in viral load between individuals to variation in the secondary infection distribution, we calculated the overdispersion in secondary infections for different combinations of “homogeneous” and “heterogeneous” contact rates and viral load trajectories. In the “homogeneous” cases, contacts were Poisson distributed with mean equal to the mean daily contacts for each time period and all individuals had the median viral load trajectory (i.e. the trajectory with the median proliferation and clearance durations and median peak Ct value), while in the “heterogeneous” cases, contacts and viral load trajectories were allowed to vary according to their full distributions.

We performed a sensitivity analysis with greater variation in viral load trajectories across individuals, by doubling the standard deviations of the distributions for the proliferation time, clearance time and peak Ct value, to test the sensitivity of the results to the degree of viral load heterogeneity. For this sensitivity analysis, we considered onward transmissions up to 88 days post exposure.

### Simulation of interventions

We assessed the hypothesis that if those with the highest viral loads are most likely to cause superspreading events, and LFTs are most sensitive for those with the highest viral loads, then lateral flow testing should be able to prevent superspreading events, which we defined as infecting over 10 contacts. We determined the probability of testing positive on an LFT at a given viral load, *p*_test_, by fitting a logistic regression model to data on LFT outcomes for isolates with different viral loads [14]. Detection was then modelled via a Bernoulli draw with this detection probability, that is,

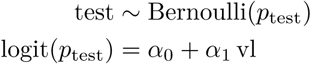

where *α*_0_ = *−*9.1 (95% CI: -13.3, 5.0) and *α*_1_ = 1.8 (95% CI: 1.1, 2.6) were the estimated coefficients from the logistic regression. We then modelled index cases testing regularly with LFTs at different frequencies (every 1, 3, and 7 days) or only before events of a certain size (before meeting >10, >20, >50 others) to determine the comparative effectiveness of rapid testing to control transmission. We assumed that a certain proportion of individuals self-isolated at home upon first testing positive, such that their non-household contacts were decreased to zero after the date of the positive test, but their number of household contacts was unchanged. The remaining individuals were assumed to continue making contacts as normal. We varied the proportion of individuals who self-isolated upon testing positive from 0-100% to model varying uptake/adherence. We assumed timing of testing was not affected by symptom onset and intensity, as we wished to model regular and event-triggered testing regardless of symptoms. We considered three indicative timepoints before (BBC Pandemic, 2018) and during the pandemic (1st lockdown (March-June 2020) and the reopening of schools (September 2020)). We reran the analysis for regular lateral flow testing with Poisson-distributed contacts instead of overdispersed contacts to look at the effect of contact heterogeneity on the impact of regular testing.

The code and data for this study can be found at https://github.com/bquilty25/superspreading_testing.

## Results

### Changes in the distribution of social contacts with changes in the intensity of restrictions

The age distribution of all participants in the BBC Pandemic survey and of the participants in each of the nine periods of the CoMix survey is shown in Figure S3. Relative to the mid-2020 age distribution of the UK population, children and older adults (70+ years) are under-represented in the BBC Pandemic data. The age distribution of CoMix participants is representative of the general population (with slight under-representation of children *≤* 4 years and over-representation of adults aged 50-69 years) and remains fairly stable across the different time periods.

Figure 2A and Table 1 show the proportions of survey participants in the UK reporting more than a certain number of contacts (5, 10, 20, 50, 100 and 200) in the previous day over time, both pre-pandemic (BBC Pandemic survey) and from the CoMix survey for the nine time periods during the survey with different levels of restrictions. The percentage of individuals reporting more than 20 contacts in a day was substantially lower during the pandemic compared to the pre-pandemic period (when it was 13.7%), varying from a low of 0.4% during the first lockdown from March to June 2020 to a peak of 6.1% in September 2020, when restrictions were most relaxed and schools reopened. The percentage of individuals reporting over 100 and 200 contacts was lower in the pre-pandemic BBC Pandemic contact survey compared to periods of relaxed restrictions due to differences in the reporting of high contact events. Overall, people reporting more than 50, 100, or 200 contacts made up less than 3% of the total CoMix survey sample.

**Figure 2.**
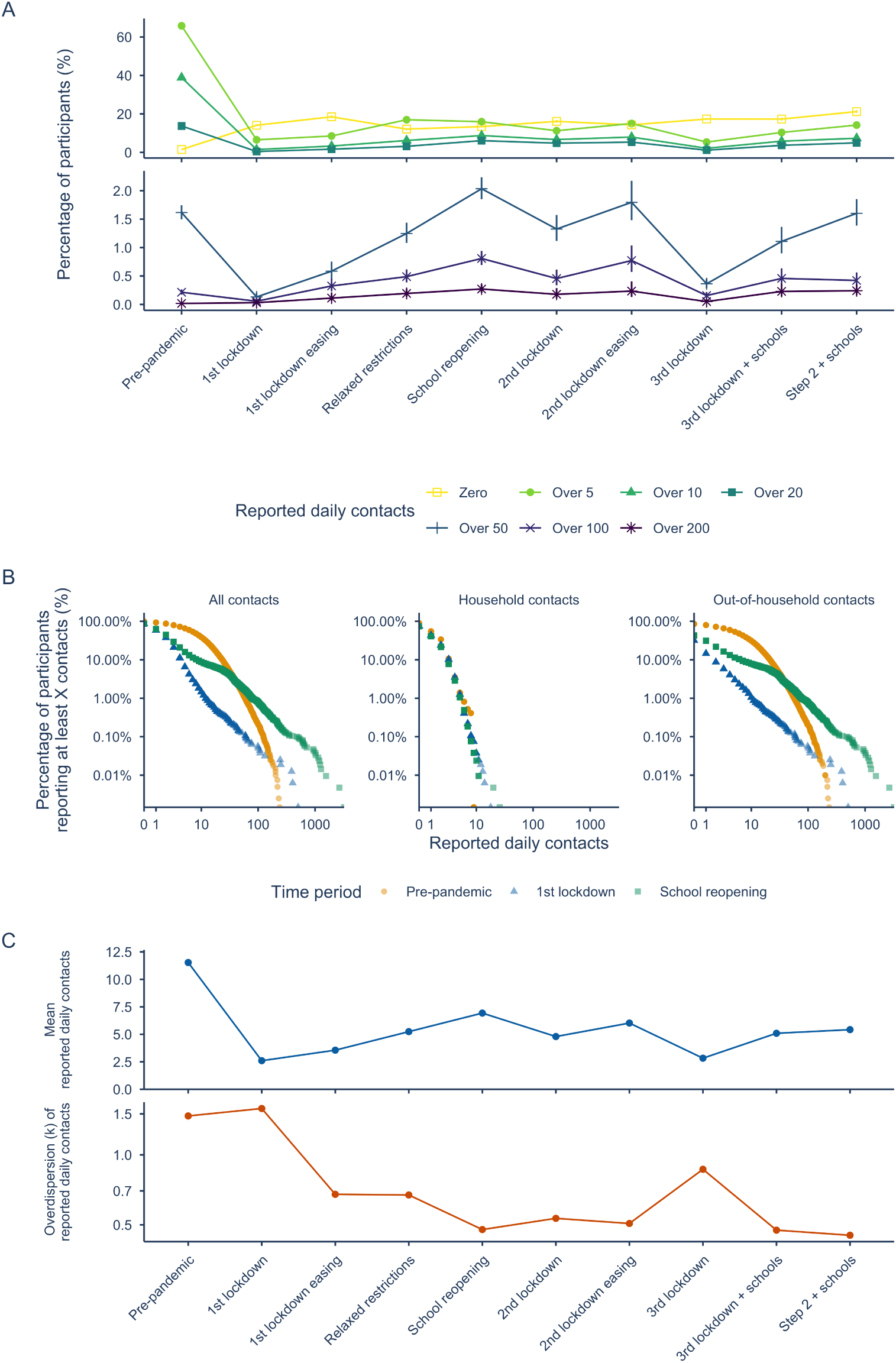
Changes in the distribution of social contacts in the UK from pre-pandemic to May 2021. A. Percentage of participants in the BBC Pandemic and CoMix contact surveys reporting zero, over 5, 10, 20, 50, 100, and 200 daily contacts in 2018 (pre-pandemic) and nine time periods during the pandemic in the UK between March 2020 and May 2021. Median and 95% binomial confidence intervals shown. B. Distribution of the number of reported daily contacts for three indicative timepoints before and during the pandemic in the UK in 2020. C. Mean and overdispersion parameter, *k*, of a negative binomial distribution fitted to contacts during the different time periods. The heavier tail in the contact distribution for the School reopening period in panel B compared with the Pre-pandemic period is likely due to mass contacts not being recorded in the BBC Pandemic contact survey, which may have truncated the contact distribution. See Figure S4 for the BBC Pandemic contact distribution with an imputed heavier tail, which is used to perform a sensitivity analysis.

Different pandemic time periods are based on lockdowns and different levels of restrictions.

*N* is the number of survey responses in each time period.

The estimated mean and overdispersion parameter *k*, with lower values indicating more overdispersion in the distribution, of numbers of daily contacts in the UK were lower than that observed pre-pandemic, with mean daily contacts averaging 4.7 during the pandemic compared to 11.5 pre-pandemic, and *k* averaging 0.7 during the pandemic compared to 1.5 pre-pandemic, indicating individuals having on average lower, but more varied, numbers of daily contacts. These values also changed considerably across the different periods of restrictions in a similar pattern to the proportion of participants with high numbers of contacts, with the mean number of daily contacts ranging from 2.6 during the first lockdown to 6.9 when schools reopened in September 2020, with a similar drop during the third lockdown, but less of a drop during the second lockdown when schools remained open (Figure 2C). The overdispersion in contacts, *k*, was similar pre-pandemic and during the first lockdown (1.5 and 1.6 respectively) then became lower, indicating more variability, following the easing of the first lockdown, with *k* between 0.5 and 0.7 in the remaining 2020 periods (Figure 2C). Stratifying contacts by household and out-of-household revealed contact rates within the household remained stable throughout, with changing non-household contact rates driving much of the variation in the mean and *k* of daily contacts over the course of the pandemic (Figure S5). Both the mean and *k* of out-of-household daily contacts remained lower than pre-pandemic contact rates. Contact durations in CoMix differed significantly between household and non-household contacts, with a median duration of 30 minutes (IQR: 5 – 180 minutes) for out-of-household contacts compared to 480 minutes (8 hours) (IQR: 180 – 1080 minutes) for household contacts (Figure S1). Figure S6, in which contacts are stratified by age as well household/out-of-household, shows that both household and out-of-household contacts in children (<18 years) remained higher than those in adults throughout 2020 and early 2021, with significant increases in out-of-household contacts of children during periods when schools were open. Contact levels were similar for males and females across all time periods.

### Variation in viral load and infectivity over the course of infection

Analysis by Kissler et al. [13] of densely sampled viral load trajectories in individuals infected with SARS-CoV-2 suggested substantial variation between individuals in the duration of proliferation and clearance phases of infection, and lowest Ct value, inversely correlated with peak viral load (Figure 3A). Mapping viral load to infectivity via a logistic function based on viral load and culture from Pickering et al. [14] (Figure 3B), calculating the probability of infectivity by day (Figure 3C), then integrating under the infectivity curve to calculate total infectious virus shed, reproduces substantial heterogeneity in individual infectiousness as reported by Ke et al. [6]. We found an 73-fold difference in individual-level infectivity between the 2.5% and 97.5% percentiles of the distribution of the area under the infectivity curve (0.08 and 5.74 arbitrary units, respectively). Fitting a Gamma distribution to this distribution gave a shape parameter of 1.42 (1.6 for Ke et al.). A substantial proportion (19.4%) of individuals were estimated to be infectious for zero days, but the remainder were infectious for at least one day (Figure 3). Across all individuals, the median duration of infectiousness around peak viral load, that is, having high enough viral load to cause infections through culturable virus, was 2 days (95% prediction interval: 0, 6 days). The majority of individuals passed through at least one day of high infectiousness (peak culture probability greater than 0.6 for 68% of individuals, greater than 0.8 for 53%), and were therefore theoretically capable of causing superspreading events given sufficient contacts of sufficient duration. However, the percentage of infected individuals who can actually initiate a superspreading event is much lower than 50% because the time window for this level of infectivity is so narrow that it must coincide perfectly with a high number of contacts of sufficient duration for a high number of infections to occur.

**Figure 3.**
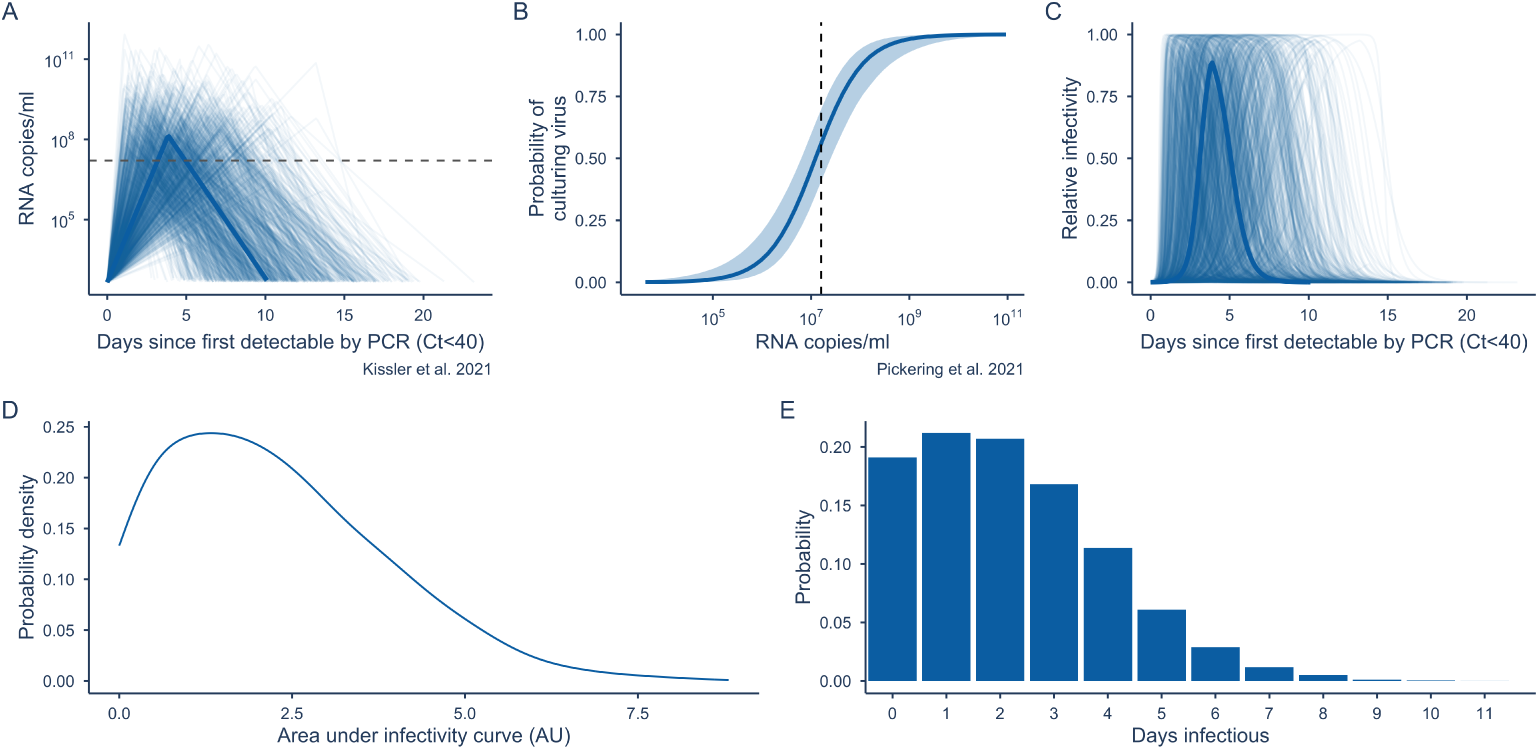
Variation in viral load progression and infectivity. A. Individual-level viral load trajectories. B. Logistic model for probability of culturing virus given a certain viral load. C. Relative infectivity over time. D. Area under the infectivity curve (in arbitrary units, AU). E. Distribution of the number of days for which individuals were infectious. Solid lines in A and C represent median viral load and infectivity trajectories. The dashed line in A and B represents the RNA copies per ml required for a 50% probability of culturing virus or infectiousness. The shaded band in B represents the uncertainty in the probability of culturing virus given a certain viral load from the fitted logistic regression model from Pickering et al. Figure S7 shows the difference in viral load trajectories and infectivity for the sensitivity analysis with double the standard deviation in the viral load trajectory parameters.

### Predicting transmission dynamics from heterogeneity in contact rates and viral load progression

Simulating secondary infections using the stochastic model described above, we estimated the overdispersion, *k*, in the initial secondary infection distribution as 0.48 (95% CI 0.47 – 0.51) (Figure 4), in line with other estimates [18, 19]. During the first national lockdown, *R* was estimated at 0.37 (95% CI 0.35 – 0.41) and *k* at 0.25 (95% CI 0.21 – 0.29). *R* rose as lockdown eased (*R* = 0.66, 95% CI 0.56 – 0.8) and restrictions were relaxed in the summer of 2020 (*R* = 1, 95% CI 0.92 – 1.1) and again as schools reopened in the autumn (*R* = 1.5, 95% CI 1.3 – 1.6). *R* then decreased again during the second lockdown to 1 (95% CI 0.92 – 1.2). As with contacts, *k* reduced as the pandemic began, indicating more variability in the secondary infection distribution, and varied between 0.12 and 0.17 after the first lockdown up to the end of 2020. A sensitivity analysis imputing a heavier tail for the BBC Pandemic contact distribution based on the tails of contact distributions from relaxed restriction periods during the CoMix survey led to a small increase in *R*_0_ from 2.5 (95% CI 2.4 – 2.6) to 2.8 (95% CI 2.6 – 3) and a small reduction in *k* from 0.48 (95% CI 0.46 – 0.51) to 0.44 (95% CI 0.4 – 0.49) (Table S1).

**Figure 4.**
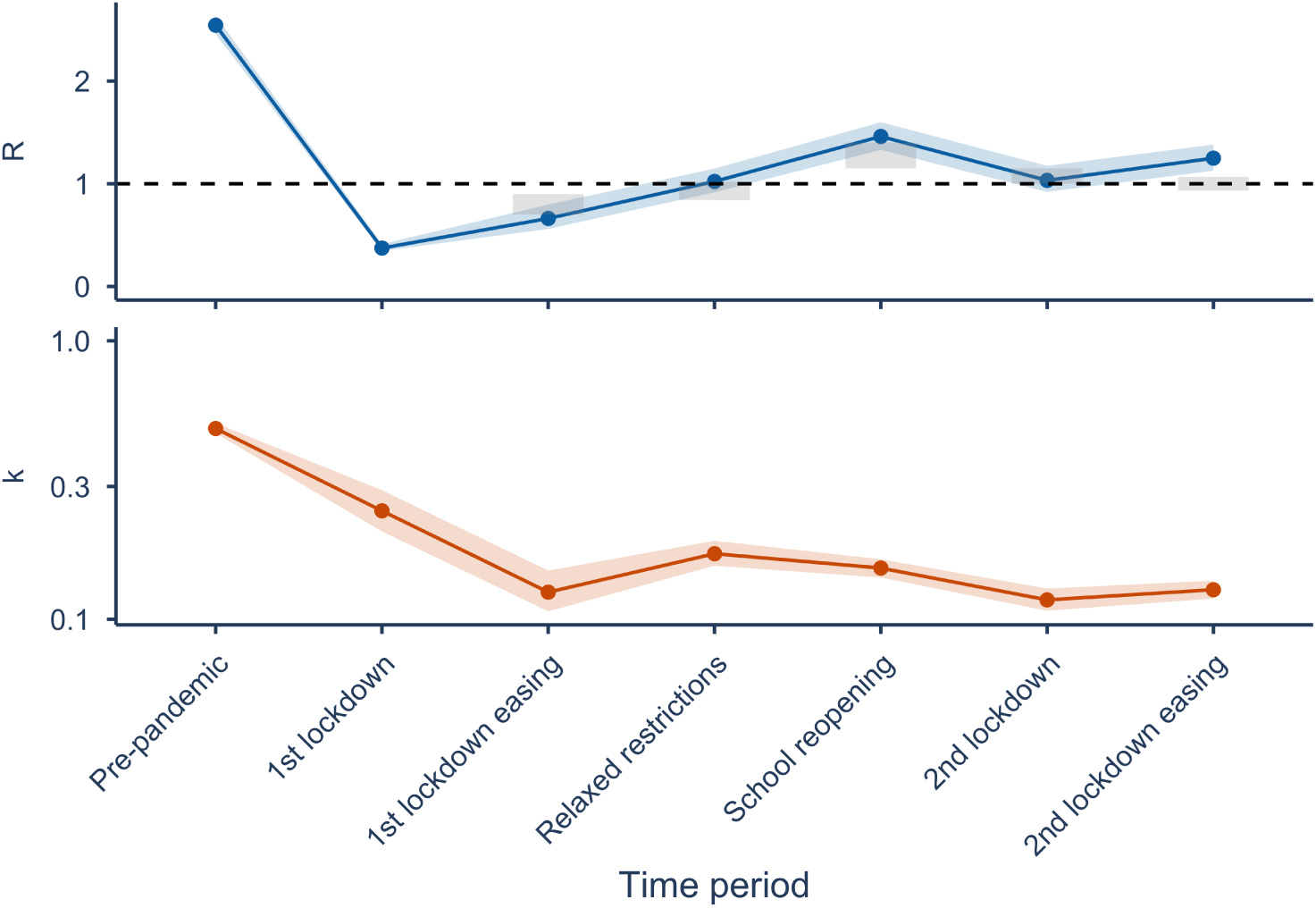
Estimates of the mean and dispersion of negative binomial distributions fitted to simulated secondary infection distributions by time period in 2020. The dispersion parameter *k* gives an indication of the variation in the secondary infection distribution, with smaller values corresponding to greater variation and values less than 1 corresponding to very large variation. Grey boxes in the *R* plot show the average upper and lower bounds (90% confidence interval) of the consensus estimates published by the Scientific Pandemic Influenza group on Modelling in the UK for the specified time periods. Shaded bands show 95% bootstrap confidence intervals for the *R* and *k* estimates.

### Contribution to heterogeneity in secondary infections due to variation in contact rates and viral load progression

If both contact rates and viral load trajectories are treated as homogeneous, with Poisson-distributed contacts and no variation in viral load trajectory between individuals, then the secondary infection distribution is approximately Poisson with equal mean and variance, corresponding to very large values of *k*. If contacts are Poisson distributed but viral load trajectories are allowed to vary between individuals, *k* is 1.5 – 1.9. If viral load is fixed at the median trajectory, but contacts are overdispersed, values of *k* are lower (0.11 – 0.74) and approximately equal to that of overdispersed contacts and variable viral load trajectories (0.11 – 0.51), indicating that overdispersion in numbers of daily contacts, as the denominator in the infection process, contributes more to superspreading, meaning overdispersion in the secondary infection distribution, than variable viral load trajectories (Figure 5). The two overdispersed-contact scenarios, equal and variable viral load, differ only pre-pandemic (*k* = 0.74 versus 0.51). This is due to individuals in the upper tail of infectiousness generating disproportionately many secondary infections when contacts are plentiful, as a result of the right-skewed area-under-the-curve distribution, exceeding an 73-fold range (Figure 3D). During restriction periods this difference disappears, as compressed contact rates create a ceiling on secondary cases regardless of viral load, and the proportion infecting zero others and more than ten others converges across the two scenarios from the first lockdown onwards (Figure 5). In the sensitivity analysis with greater variation in individuals’ viral load trajectories, with double the standard deviation in the viral load trajectory parameters, the *k* estimates for the scenario with Poisson contacts (0.76 – 0.92) and variable viral loads were closer to those for overdispersed contacts and variable viral loads (0.08 – 0.33), but still higher (Figure S8). The finding that contact heterogeneity plays a greater role in overdispersion in secondary infections than viral load heterogeneity therefore appears to be robust to much greater variation in peak viral load and duration of viral shedding than found in the cohort in Kissler et al. [13].

**Figure 5.**
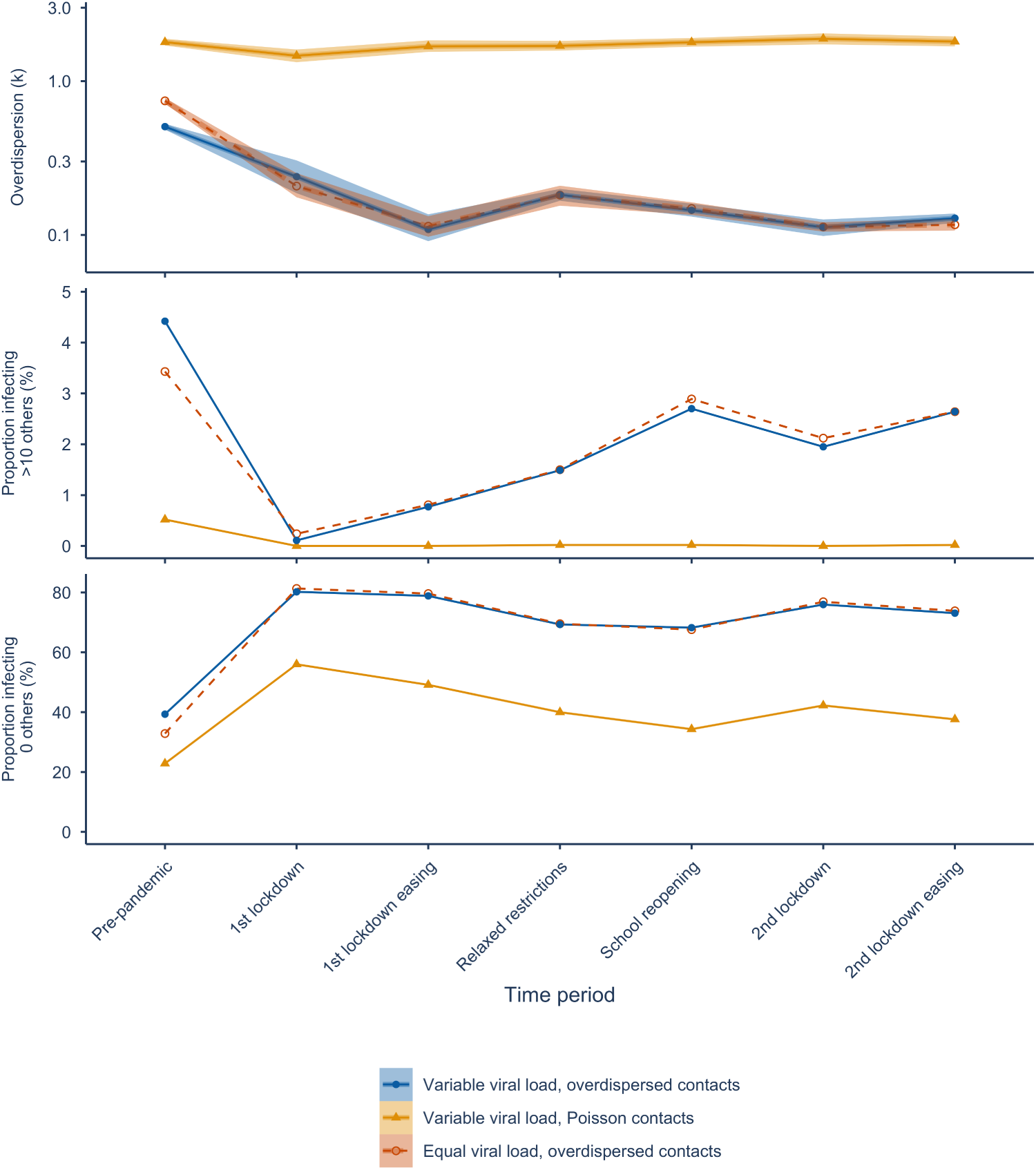
Estimates of the dispersion of negative binomial distributions fitted to secondary infection distributions by period, with different combinations of homogeneous and heterogeneous contact rates and viral load trajectories, along with the proportion of individuals infecting zero others and more than ten others. The dispersion parameter *k* gives an indication of the variation in numbers of secondary infections, with smaller values corresponding to greater variation and values less than 1 corresponding to very large variation. Values of *k* for equal viral load and equal contacts are not shown because they are very large, meaning the numbers of secondary infections are approximately Poisson distributed. Shaded bands show 95% bootstrap confidence intervals. The sensitivity analysis with greater variation in viral loads is shown in Figure S8.

### Impact of rapid testing (regular testing vs. pre-event testing)

For pre-pandemic levels of contacts, uptake of regular LFT testing must exceed 70% to reduce *R* below 1 if testing daily and must be above 80% if testing every 3 days, but even 100% uptake will not reduce *R* below 1 if testing weekly (Figure 6A). For lockdown levels of contacts, testing does not appreciably reduce *R* below its already low level. For contact rates equivalent to those under relaxed restrictions with schools open, testing every 3 days will reduce *R* below 1 if uptake exceeds 70% (Figure 6A). Pre-event testing acts similarly, with high uptake (> 90%) and testing for events with more than 10 others necessary to reduce *R* below 1 for pre-pandemic levels of contact, but only testing when attending events with more than 20 others and 60% adherence required during relaxed restrictions with schools open (Figure 6B). Both regular testing and pre-event testing reduce the rate of superspreading as defined as the proportion of individuals infecting over 10 others. However, there is also an increase in the proportion of individuals that infect no one. In some cases, this results in a decrease in *k* (greater overdispersion in the secondary infection distribution) (Figure 6). From the sensitivity analysis comparing the impact of regular testing with and without overdispersion in contacts (Figure S9), we can see that ignoring overdispersion in contacts reduces the estimated impact of testing in terms of reducing *R* and decreasing the proportion of superspreading events. This demonstrates the importance of accounting for contact overdispersion when estimating the impact of interventions on secondary infections.

**Figure 6.**
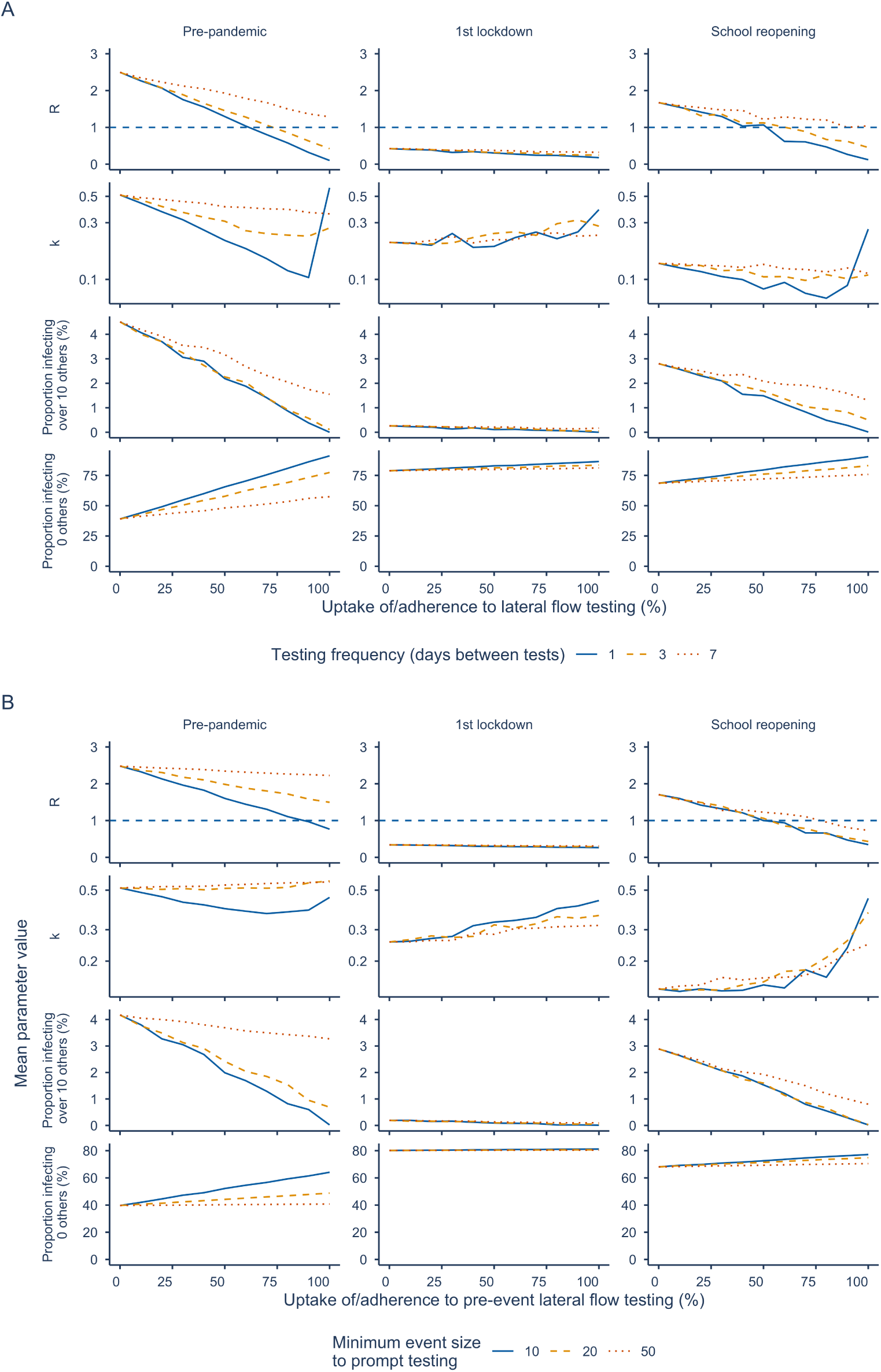
Effect of regular and pre-event lateral flow testing on *R*, *k*, and the proportion infecting over 10 or 0 others for varying levels of uptake or adherence (horizontal axes) and background contact rates (columns). A. Regular testing. B. Pre-event testing.

## Discussion

We have estimated the secondary infection distribution for SARS-CoV-2 over the course of the pandemic in the UK in 2020 using a stochastic model incorporating viral load trajectories and reported daily contacts. Our estimates of the mean number of secondary infections are consistent with contemporaneous estimates (Figure 4). Daily contact rates became both lower on average, from 11.5 per day to 4.7 per day, and more overdispersed during the pandemic in the UK in 2020 as some individuals maintained high contact rates, such as essential workers, while others had their number of daily contacts reduced to zero, such as those working from home. This drove a decrease in the mean number of secondary infections per infected individual, the reproduction number, but an increase in the dispersion in the number of secondary infections. Our estimates of the degree of overdispersion in the secondary infection distribution are similar to those reported by other authors, which are mostly between 0.3 and 0.6 [18, 19], though lower estimates of around 0.1 have been reported from outbreak cluster size data [1].

Our analysis shows that changes in the mean and overdispersion of daily numbers of contacts over time (Figure 2C) can be used to estimate dynamic shifts in the mean and overdispersion of the reproduction number (Figure 4). This motivates collecting contact data during epidemics, as contact overdispersion could provide a potential indicator of superspreading risk in real-time, along with the effective reproduction number. Changes in contact overdispersion would indicate when and where the population is at risk of superspreading events, and enable implementation of targeted, rather than blanket, restrictions on contact levels to limit outbreaks. However, ground-truth data on secondary infection distributions, e.g. from contact tracing studies, would first be required to validate estimates of the mean and overdispersion of the secondary infection distribution.

Contact heterogeneity was found to contribute more to superspreading than heterogeneity in viral load, as the number of contacts an individual makes places an upper limit on the number of people they can infect, and, despite some individuals having much higher infectious potential than others, most infected individuals pass through a high viral load period, with 68% having a peak culture probability greater than 0.6 and 53% greater than 0.8. Viral load heterogeneity does still contribute significantly to heterogeneity in transmission since viral load varies by orders of magnitude over time within individuals and between individuals (Figure 5). This indicates that superspreading is more a result of between-person contact variability and within-person infectiousness variability than between-person infectiousness variability. Hence if reductions in contact rates can be targeted specifically at individuals ahead of or during their window of high infectivity, for example by encouraging contacts of cases to undergo regular self-testing and self-isolation upon a positive rapid test result [20], then this may result in reductions in transmission while minimising the burden of quarantine. Other studies investigating the superspreading nature of SARS-CoV-2 have come to similar conclusions [4, 6, 3, 21, 22, 23]. However, as far as we are aware our study is the first to estimate the relative roles of contact and viral load heterogeneity and the impact of changes in contact heterogeneity using real-world contact distributions from before and during the pandemic and data on viral load trajectories and infectiousness [14]. Our use of reported heterogeneity in viral load trajectories [13] also contributes to a wider estimated distribution for the number of days individuals are likely infectious, which closely matches empirical daily sampling [6] and human challenge studies [5]. Our approach of using culture positivity as a direct virological proxy for infectiousness is complementary to that of Marc et al. [24], who inferred the viral-load-infectiousness relationship from observed transmission outcomes and similarly found that viral load alone does not fully explain transmission heterogeneity, with a residual random effect standard deviation of 85%. Consistent with our findings, Kendall et al. [25] found that contact rates explained more daily variability in *R*(*t*) than per-contact transmissibility (46% versus 30%) using NHS COVID-19 app data that tracked CoMix closely; Ferretti et al. [26] similarly found that contact duration drove transmission risk more than proximity, supporting the use of duration-weighted contacts in our model.

Like others [6, 23, 27], we hypothesised that lateral flow testing, by detecting individuals with high viral loads when they were most infectious, would reduce transmission through reducing the potential for superspreading. This manifested as a decrease in the proportion infecting over 10 others and a substantial increase in the proportion infecting zero others. This, perhaps counter-intuitively, resulted in a decrease in *k* in some cases, as the relative increase in those infecting zero others exceeded the decrease in those infecting many others, here defined as over 10 others. Hence, assessment of superspreading solely via the metric of the overdispersion parameter *k* may conceal changes in both the upper and lower tail of the secondary infection distribution. Both regular testing and pre-event testing were effective in reducing *R* given high enough frequency or a low enough event size threshold, respectively, as long as uptake or adherence was high. Testing had the highest relative impact on transmission when contact rates were high, for example at pre-pandemic levels, as there were more potentially preventable exposures, meaning rapid testing could reduce *R* below the growth threshold of 1 while otherwise maintaining relatively normal contact rates. In contrast, testing during lockdown would have less impact as *R* was already below 1. Having everyone in the population test every 3 days would bring *R* below 1 and be approximately equivalent in terms of impact on secondary infections to having people test only before events of minimum size 10 for pre-pandemic contact levels. This indicates that rapid testing could be an effective, minimally disruptive intervention to reduce transmission if uptake or adherence could be maximised through incentivising use. Our findings align with those of Ke et al. [6], who estimated using a within-host model of infection that daily antigen testing could reduce infectiousness by 85% but that impact reduced significantly with decreasing testing frequency. However, our analysis goes further by estimating the impact of different rapid testing frequencies on the reproduction number accounting for the full distribution of daily contacts. Hart et al. [23] similarly found that daily lateral flow testing could significantly reduce numbers of secondary infections, preventing around 70% of infections assuming complete isolation of infected individuals following detection, although they used a full multiscale model of within-host dynamics and transmission and focused on the Omicron variant, for which the effectiveness of testing may be different due to differences in within-host dynamics and pre-existing immunity.

In their analysis of the CoMix data, Gimma et al. [11] found that school-aged children consistently had higher numbers of contacts across both lockdowns and periods of relaxed restrictions than working-age adults, who in turn had higher numbers of contacts than older adults (60+ years). Contact levels also varied with employment status, with part-time-employed participants having more contacts than full-time or self-employed participants. Another analysis of the CoMix data that focused on individuals with high numbers of contacts [28] identified that most reports of high numbers of contacts came from work and school settings and tended to be reported by participants in certain professions (teaching, retail, administration, construction, medicine, social work, hospitality). Some of this work is considered essential, and keeping schools open where possible is a high priority, so opportunities for reducing these contacts may be limited. This highlights the potential value of regular rapid testing as a way of preventing superspreading even when some individuals have many contacts.

Our analysis has some limitations. The CoMix contact survey was designed to be comparable to previous contact surveys in the UK, namely the BBC Pandemic [7, 8, 9] and POLYMOD [10] surveys. However, previous surveys required participants to list contacts individually to include other information such as contact age, sex, and occupation, meaning that it was difficult to include mass contacts such as those made at a large gathering, thus truncating the true contact distribution (Figure 2B). From 18 May 2020 the CoMix survey introduced the option to record mass contacts as a count rather than listing each contactee in a separate entry. This means pre-pandemic and early-pandemic contact distributions are not directly comparable to those conducted later. However, we conducted a sensitivity analysis to check how this could impact our estimates of numbers of secondary infections, by imputing a heavier tail for the BBC Pandemic contact distribution based on the tails of the contact distributions during periods of relaxed restrictions in the CoMix survey, and observed only a small increase in *R*_0_ and a small reduction in *k*. Additionally, variation between participants in how direct contacts are interpreted and reported could contribute to artificial overdispersion in the contact distribution and hence in our estimates of secondary infection overdispersion. Another limitation of the BBC Pandemic contact data is that working-age adults were overrepresented among the participants, while children under 13 years were excluded (Figure S3). It is possible that this may have led to some bias in the reported contact distributions. However, the similarity of contact patterns observed in the BBC Pandemic and UK POLYMOD datasets [8] suggests that the BBC Pandemic data is broadly representative of pre-pandemic contact patterns in the whole UK population.

We used data on viral loads over time from a detailed study of individuals in the National Basketball Association cohort in Kissler et al. [13], which is unlikely to be representative of the full variation in viral load trajectories in the general population, since the cohort consisted mostly of young, healthy, male professional athletes. Our analysis could therefore underestimate the variability in secondary infections due to variability in viral load with factors such as age, for which there is some evidence of an effect on infectiousness [6], co-morbidities, and sex. In particular, the NBA cohort does not include individuals with very long infections. Ghafari et al. [29], for instance, found that some individuals in the Office for National Statistics COVID-19 Infection Survey in the UK had persistent infections lasting more than 60 days. However, they constituted only 0.1 to 0.5% of all infections, with PCR-derived viral loads around 100-fold lower than at symptom onset, substantially reducing their transmission potential. Furthermore, PCR measurements were only taken from ONS infection survey participants weekly for a month and then monthly up to a year, whereas measurements were near daily in the NBA cohort and crucially captured the early stage of infection, and therefore permit much higher resolution modelling of onward transmission. We conducted a sensitivity analysis with much greater variation in viral load trajectories to account for greater variation in the general population and found that it did not qualitatively change our conclusion that contact heterogeneity remains the primary driver of heterogeneity in transmission.

We assume that the probability of shedding infectious virus is equal to the probability of culturing virus, but the exact relationship between viral culture positivity and infectiousness is not known [30], so our results must be interpreted with this limitation in mind. Furthermore, the specific proportions of infectious individuals we report depend on the fitted logistic dose-response curve between viral load and culture probability; alternative functional forms would yield different estimates. A structural asymmetry in our comparison is that contact heterogeneity is measured directly from survey data, whereas infectiousness heterogeneity is captured only via viral load as a proxy. Sources of between-individual variation in infectiousness that are decoupled from within-host viral dynamics, such as variation in respiratory droplet production, speaking volume, or interpersonal distance preferences, are not captured, and if substantial would increase the true contribution of infectiousness heterogeneity relative to our estimates. We partially address this by drawing the logistic regression coefficients linking viral load to infectiousness for each individual from the estimated covariance distribution, and by examining robustness in a sensitivity analysis with twice the standard deviation in viral load trajectory parameters (Figure S8); however, these measures do not capture variation in transmission efficiency that is entirely independent of viral load. We also do not account for correlation of the timing of peak viral load and infectiousness with symptom onset and intensity [31], which may affect contact rates and numbers of secondary infections [24].

We focus on an index case and their infections in one other generation, which may underestimate second-order effects that considering a full contact network structure would capture [6]. For instance, we would expect individuals that acquire infection to typically have higher contact rates than the average member of the population, since having more contacts increases acquisition risk, and therefore to infect more individuals. However, incorporating this effect would require additional assumptions about how acquisition risk scales with contact number and network structure that we do not have the data to support. We do not consider other interventions that may have an additional impact on *R*, such as vaccination, contact tracing, or self-isolation upon symptom onset. We also do not consider the impact of variants with increased transmissibility, and hence limit our analysis to 2020 before the widespread emergence of variants of concern such as Alpha, Delta, and Omicron.

We assumed within-household contacts for each individual were the same each day, and sampled daily non-household contacts from repeat responses for the same survey participant, which may underestimate variation in overall daily contact rates and hence infection potential of specific individuals. We assume that self-isolating individuals are unable to fully self-isolate from their household members as reported by the majority of those surveyed by the ONS in England in April 2021 [32]; further decreases in *R* may be possible if self-isolating individuals isolate themselves from household members.

There is some evidence that heterogeneity in secondary infections has changed as new SARS-CoV-2 variants of concern have emerged and population immunity has increased. Studies examining heterogeneity in SARS-CoV-2 transmission over time in Japan and Europe have found that overdispersion remained after the emergence of Alpha, Delta, and Omicron, with *k* generally being below 0.5 but varying somewhat across waves and being higher, that is, less overdispersed transmission, during the Omicron wave [33, 34]. Meta-analyses of literature estimates of *k* have shown a wide range of estimates across settings, clusters and variants, but no clear trend in *k* over time [35, 36], though some studies suggested slight increases in *k* with the emergence of Delta. A modelling study has suggested that non-pharmaceutical interventions such as lockdowns have imposed selective pressure on SARS-CoV-2 towards more homogeneous transmission, corresponding to higher *k* [37]. Together, these studies suggest that while overdispersion may be weakening with widespread immunity and new variants of concern, it remains a feature of SARS-CoV-2 transmission.

Thus, while our analysis is restricted to 2020, our findings are still relevant in the current era of endemic COVID-19 and to other pathogens that exhibit heterogeneous transmission, including other SARS coronaviruses, Ebola, measles, influenza, RSV, and adenoviruses [3, 38, 39]. Moreover, the question of the relative contribution of variation in numbers of contacts and variation in infectiousness to heterogeneity in transmission is a universal one for infectious diseases. By demonstrating that superspreading potential is closely linked to the overdispersion of social contacts and the timing of peak infectiousness, we have shown the importance of monitoring not only the mean effective reproduction number but also its variability. Measurement of contact distributions thus offers a real-time surveillance tool for identifying superspreading risk, supporting pandemic preparedness through enabling targeted interventions against emerging pathogens. The modelling framework is generalisable: provided densely sampled pathogen load trajectories and pathogen-specific culture or test sensitivity data are available to characterise infectiousness, and a contact survey with duration information is available to characterise the contact distribution, the same approach can be applied to estimate heterogeneity in the secondary infection distribution for other pathogens.

Our results suggest superspreading for SARS-CoV-2 can be best explained as a random sample from the tail of the contact and shedding distribution: it occurs when an infected individual makes a high number of contacts during a highly infectious period lasting approximately 2 days on average, with the majority of infected individuals being sufficiently infectious, 68% with a peak culture probability over 0.6 and 53% over 0.8, for at least one day to be capable of causing a superspreading event, given they make a high number of contacts. Changes in the number of contacts observed throughout the pandemic in the CoMix contact survey were able to explain changes in the reproduction number, with contact rates becoming more heterogeneous during the pandemic given lockdowns and changes in working practices. For future pandemics, regular or pre-event lateral flow testing may be an efficient way to target individuals when most infectious and hence minimise the burden of non-pharmaceutical interventions while maximising reduction in transmission rates, provided uptake is moderate to high.

## Data Availability

All data used in the study are available online at https://github.com/bquilty25/superspreading_testing

## Supporting Information

**Figure S1 Distribution of the duration of contact for household and out-of-household contacts from the CoMix contact survey in the UK.**

**Figure S2 Correlation between number of contacts and mean contact duration for CoMix survey participants from March 2020 to May 2021.**

**Figure S3 Age distribution of participants in the BBC Pandemic (Pre-pandemic) contact survey and each of the nine time periods from the CoMix contact survey from March 2020 to May 2021 relative to the mid-2020 UK age distribution [40].**

**Figure S4 Contact distributions for three indicative timepoints before and during the pandemic in the UK with an imputed heavier tail in the Pre-pandemic (BBC Pandemic) contact distribution (cf. Figure 2B).** The heavier tail was imputed from the tails of contact distributions during periods of relaxed restrictions in the CoMix survey. We conducted a sensitivity analysis with this distribution to determine the effect of the heavier tail on *R*_0_ and *k*.

**Figure S5 Distribution of the number of reported daily contacts for all time periods before (BBC Pandemic contact survey) and during (CoMix contact survey) the pandemic in the UK between March 2020 and May 2021.**

**Figure S6 Contact distributions by age and gender across the nine time periods from the CoMix contact survey from March 2020 to May 2021.**

**Figure S7 Comparison of viral load trajectories and infectivity for the sensitivity analysis with double the standard deviation in viral load trajectory parameters and those used in the main analysis.** A. Individual-level viral load trajectories. B. Logistic model for probability of culturing virus given a certain viral load. C. Relative infectivity over time. D. Area under the infectivity curve, in arbitrary units. E. Distribution of the number of days for which individuals were infectious. Solid lines in A and C represent median viral load and infectivity trajectories. The dashed line in A and B represents the RNA copies per ml required for a 50% probability of culturing virus or infectiousness. The shaded band in B represents the uncertainty in the probability of culturing virus given a certain viral load from the fitted logistic regression model from Pickering et al.

**Figure S8 Sensitivity of estimates of dispersion of the secondary infection distribution in different time periods to the degree of variation in viral load trajectories.** Left: standard deviation in distributions for viral load trajectory parameters from Kissler et al. Right: standard deviation of viral load parameters doubled.

**Figure S9 Comparison of the impact of regular lateral flow testing on** *R*, *k***, and the proportion infecting over 10 or 0 others with Poisson-distributed contacts and overdispersed contacts, for varying levels of uptake or adherence and background contact rates.**

**Table S1 Sensitivity analysis of the effect on** *R* **and** *k* **when imputing a heavier tail of the contact distribution for the pre-pandemic BBC Pandemic contact survey, which lacked the ability to record raw counts of high numbers of contacts.**

**Table S2 Sensitivity analysis of the effect on the mean and dispersion in contact rates comparing a negative binomial distribution to a zero-inflated negative binomial distribution.**

